# Treatment and Evaluations among Infants Exposed to Syphilis in Utero, 6 U.S. States, 2018–2021

**DOI:** 10.1101/2025.09.02.25334918

**Authors:** Mundayi V. Nlandu, Elizabeth L. Lewis, Jeffrey M. Carlson, Ellen Martinson, Keivon Hobeheidar, Abigael Gardere, Teri’ Willabus, Deborah Mbotha, Nicole D. Longcore, Heather Marshall, Kevin P. O’Callaghan, Van T. Tong, Kate R. Woodworth

## Abstract

Contemporary data on treatment and evaluations of infants exposed to syphilis in utero are limited. This study analyzed population-based surveillance data to describe the management of exposed infants. Findings indicate inconsistent implementation of recommended guidelines for treatment and evaluation of infants exposed to syphilis in utero.

## Introduction

The incidence of congenital syphilis (CS) continues to rise in the United States with >740% increase from 2014 to 2023^1^, despite available preventative measures.^2,3^ Treatment and evaluations for syphilis in pregnancy and CS are complex. The 2021 Sexually Transmitted Infections (STI) Treatment Guidelines recommend treatment and evaluations for infants born to women with syphilis during pregnancy.^3^ These guidelines are based on clinical scenarios of likelihood of CS, dependent on maternal testing and treatment, as well as infant laboratory and physical exam findings.^3^ Provider surveys have found varied approaches in care of exposed infants, but data on real-world evaluation and treatment patterns are outdated or geographically limited.^4,5^ Due to subtle differences between CS categorization in the STI Treatment Guidelines and surveillance criteria in the Council of State and Territorial Epidemiologists (CSTE) CS case definition, some infants who are recommended to receive testing and treatment are not included in national syphilis surveillance data.^3,6^

Using a population-based surveillance cohort, we describe treatment patterns and evaluations by clinical scenario among infants exposed to syphilis in utero.

## Methods

The Surveillance for Emerging Threats to Mothers and Babies Network (SET-NET) collects enhanced syphilis surveillance data in select U.S. jurisdictions.^7^ Detailed methods have been previously published.^8,9^ Six states (Arizona, Georgia, Michigan, New Jersey, New York [excluding New York City], and Washington) ascertained syphilis during pregnancy through national notifiable disease reporting and linkages with vital records.^8^ Data were collected using case reports, medical records, vital records, and laboratory results. Infants born January 1, 2018–December 31, 2021 were included if the woman met the CSTE case definition for syphilis during pregnancy or infant met the probable or confirmed CSTE case definition for CS.^6^ Analysis included infants with completed birth hospitalization medical record abstraction and who were reported to SET-NET as of June 7, 2024.

Infants were categorized into clinical scenarios: 1) *confirmed or highly probable CS* defined as having a positive darkfield test OR abnormal physical examination with signs consistent of CS OR an infant nontreponemal serologic titer ≥fourfold the maternal titer at delivery; 2) *possible CS* defined as not meeting scenario 1 and inadequate or no maternal syphilis treatment in pregnancy; or 3) *CS less likely* defined as not meeting scenario 1 and adequate maternal syphilis treatment in pregnancy.^3^

STI treatment guidelines recommend infants with confirmed or highly probable CS receive 10 days of penicillin treatment, cerebrospinal fluid (CSF) evaluations, long-bone radiographs, and complete blood count with differential (CBC/D).^3^ Infants with possible CS are recommended to either have 10 days of treatment, or a single dose of penicillin provided they have a normal CSF evaluation, long-bone radiographs, and CBC/D. Infants with CS less likely are recommended to either receive one dose of penicillin or have close serologic follow-up. All infants with reactive nontreponemal titers at birth are recommended to have follow-up serologic testing.

Descriptive analyses were presented for infants by CS clinical scenarios and by birth year, gestational age, neonatal intensive care unit (NICU) admission, and NICU admission among those ≥35 weeks gestational age. Clinical scenario-based recommended evaluations included long-bone radiographs and CSF analyses. Treatment categories reported included 10 days of aqueous crystalline penicillin G (PenG) or procaine PenG (10 days of PenG), one dose of benzathine penicillin G (BPG), other treatment (other non-recommended penicillin or non-penicillin-based treatment), or no treatment. In a sub-analysis of infants with reactive nontreponemal titers at birth and follow-up data available up to 9 months, we reported frequency of follow-up serologic testing.

Analyses were conducted using R version 4.4 (R Foundation) statistical software. This activity was reviewed by CDC and conducted consistent with applicable federal law and policy (*45 C*.*F*.*R. part 46*.*102(l)(2), 42 U*.*S*.*C. Sect. 241(d); 5 U*.*S*.*C. Sect. 552a*).

## Results

A total of 1636 infants met inclusion criteria; 6 received an incomplete course of treatment with 4 to 7 days of aqueous PenG (5 with possible CS and one CS less likely), were considered unclassifiable and excluded from the analysis. Of the remaining 1630, 179 infants were categorized as confirmed or highly probable CS (11.0%), 534 (32.8%) as possible CS, and 917 (56.3%) as CS less likely (Table 1).

**Table 1.**
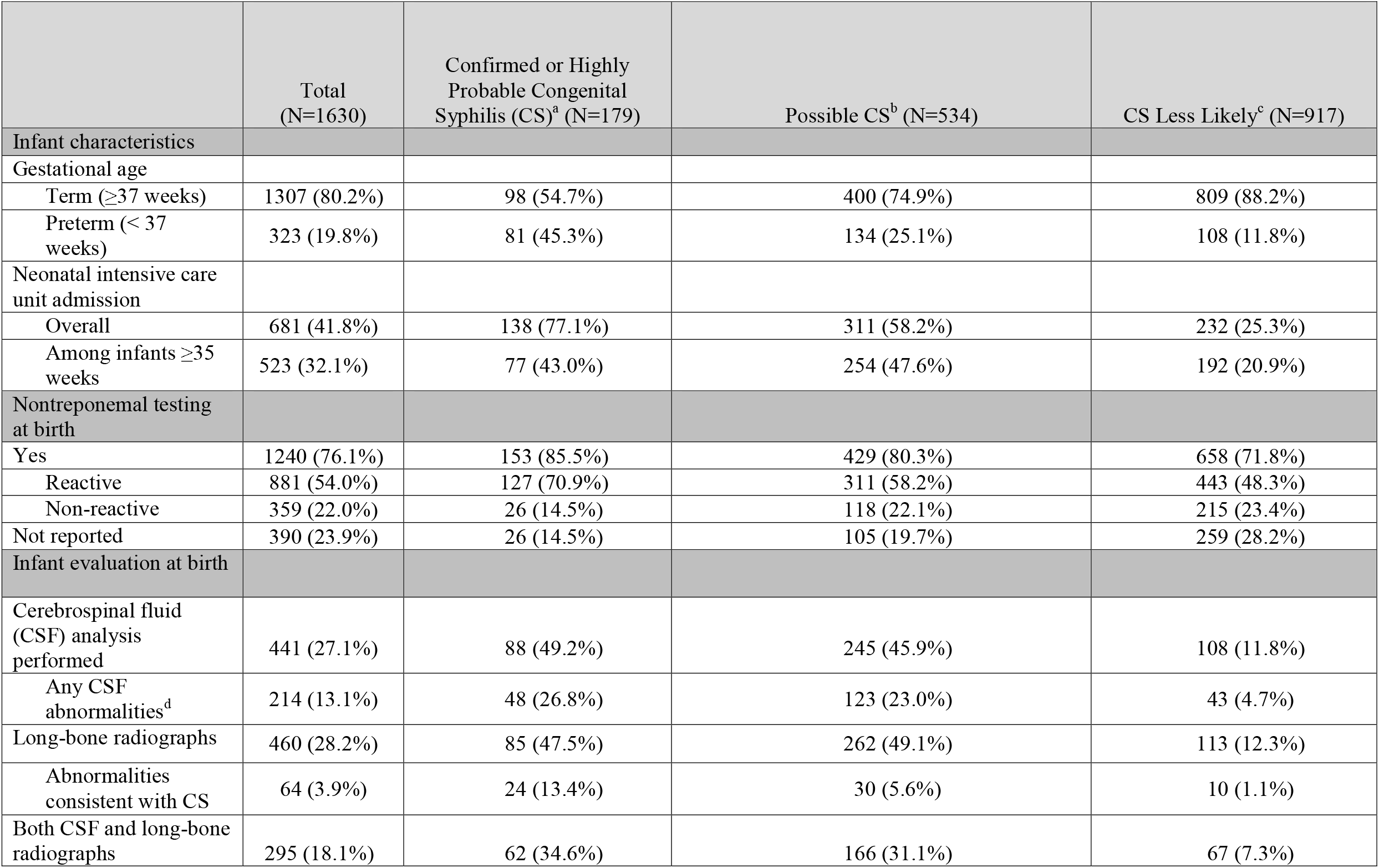
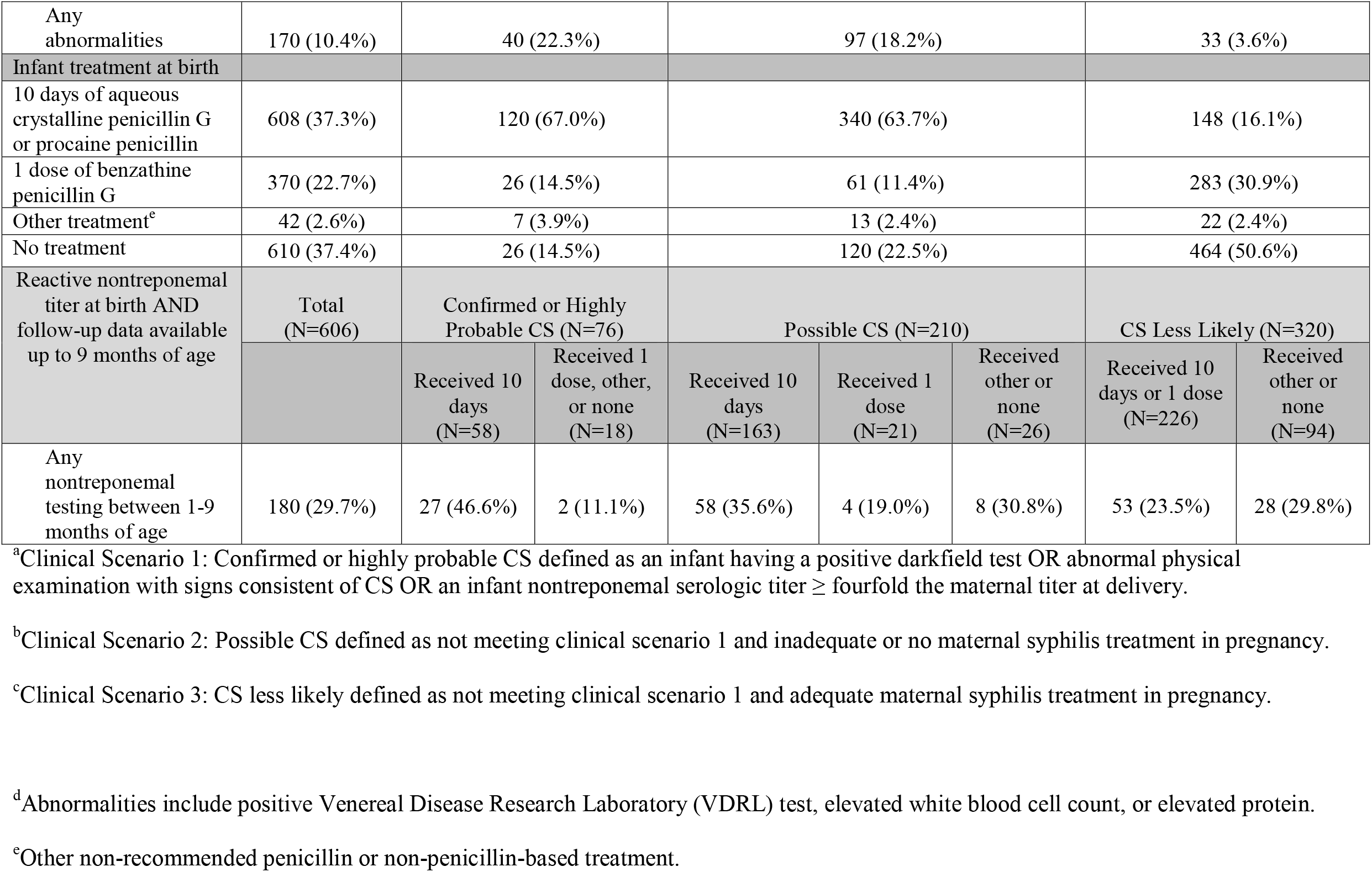
Characteristics, evaluations, and treatment among infants exposed to syphilis in utero, 6 U.S. States, 2018–2021.

Of 179 infants with confirmed or highly probable CS, 81 (45.3%) were preterm and 138 (77.1%) were admitted to a NICU. Two-thirds (67.0%) received the recommended 10 days of PenG. Twenty-six (14.5%) received one dose of BPG and 33 (18.4%) had other or no reported treatment. Long-bone radiographs were reported for 47.5% infants and CSF for 49.2%.

Of 534 infants categorized as possible CS, 134 (25.1%) were preterm and 311 (58.2%) were admitted to a NICU. The majority, 340 (63.7%) received 10 days of PenG. Sixty-one infants (11.4%) received one dose of BPG. Of those receiving one dose, only 1 (1.6%) had both normal long-bone radiographs and CSF reported by date of treatment. Other or no treatment was reported for 133 infants (24.9%).

Of 917 infants categorized as CS less likely, 148 (16.1%) received 10 days of PenG, and 283 (30.9%) received one dose of BPG. Half (53.0%) had other or no treatment reported. While not universally recommended for this scenario, 12.3% had long-bone radiographs reported and 11.8% CSF.

For the 881 infants with a reactive nontreponemal test at birth, 606 (68.8%) had medical record abstraction of a well-child visit (>28 days to <9 months of age) or laboratory data submitted after birth hospitalization. Nontreponemal testing at follow-up was reported for 38.2% of infants with confirmed or highly probable CS, 33.3% with possible CS, and 25.3% with CS less likely.

## Discussion

We observed inconsistencies of recommended treatment and evaluations across all clinical scenarios during birth hospitalization and early infancy. While most infants with confirmed or highly probable CS received some type of treatment, only two-thirds had the full 10-day recommended course of treatment. Only a third of infants in this scenario had both long-bone radiographs and CSF evaluations reported, despite those being recommended for this group. However, for this scenario. these evaluations do not impact treatment decisions and there is debate about the necessity, specifically for invasive lumbar punctures.^10^

For infants with possible CS, these evaluations do impact recommendations for length of treatment and follow-up. Most infants with possible CS were treated conservatively with the full 10 days of treatment, where long-bone radiographs and CSF, while possibly informative, are not required. Conversely, among infants who received one dose, many did not have the normal evaluations reported, as recommended for this treatment option. Also, a quarter of infants with possible CS had other or no treatment reported.

Of infants with CS less likely, 16.1% received longer length of treatment than recommended for this scenario (10 days of intravenous or intramuscular therapy). We found a high frequency (>10%) of long-bone radiographs and CSF evaluations among this group where these evaluations are not universally recommended.^3,10^

Per STI Treatment Guidelines, infants with a reactive nontreponemal test should receive follow-up examinations and testing every two to three months until the test is nonreactive.^3^ A low proportion of infants (29.7%) with reactive nontreponemal tests received follow-up nontreponemal tests across all clinical scenarios. Infants may not receive follow-up testing due to external circumstances such as limited access to care.^4^

Public health plays a critical role in the primary and secondary prevention of CS, ensuring information on maternal testing and treatment is available to providers caring for infants and providing resources and guidance regarding recommended treatment and follow-up care. Despite available guidelines, diagnoses of CS are missed and, even among infants known to be exposed, information needed to make treatment decisions may be unavailable.^2,11^ Given the recent surge of CS, providers may be unfamiliar with guidelines, particularly for infants with CS less likely. One survey conducted in 2024 showed that of 442 pediatric providers 94.1% selected to evaluate and manage infants with confirmed or highly probable CS as recommended, yet only 45.8% did the same for infants with CS less likely.^4^

This large, population-based analysis utilized surveillance data from six jurisdictions representative of areas with high CS prevalence. However, there are several limitations. First, titers were unavailable for 25.1% (364/1451) of infants classified as possible CS and CS less likely, resulting in a maternal treatment-based classification, which could lead to misclassification of infant scenario. Second, surveillance data are subject to missing data (e.g., transfers to tertiary care or loss to follow-up) which could lead to differential underreporting of treatment and evaluations. Third, these surveillance data do not capture CBC/D values, which could influence individual treatment decisions.^3^ Finally, many complex and unmeasured factors go into clinical decision making, however, our aim is to describe a broad picture of current patterns of management to identify areas to improve the standard of care for impacted infants and families.

Amid a national syphilis epidemic, these findings emphasize the need for accessible education or consultation (e.g., through the National Network of STD Clinical Prevention Training Centers)^12^, system-level supports (e.g., integrated clinical pathways), and continued collaboration between public health and clinical partners to ensure all infants exposed to syphilis in utero receive recommended care.

## Data Availability

All data produced in the present work are contained in the manuscript.

## Author Contributions

M.V.N. conceptualized the analysis, conducted data analyses, drafted the initial manuscript, interpreted data, and reviewed and revised the manuscript. K.R.W. designed the study, conceptualized the analysis, drafted the initial manuscript, interpreted data, and reviewed and revised the manuscript. E.L.L., J.M.C., K.P.O.,V.T.T provided technical assistance and subject matter expertise for data collection, analysis, and interpretation, and critically reviewed and revised the manuscript. E.M. conducted data analyses, interpreted data, and reviewed and revised the manuscript. K.H., A.G., T.W., D.M., N.D.L., H.M. coordinated and supervised data collection and reviewed and revised the manuscript. All authors approved the final manuscript as submitted and agreed to be accountable for all aspects of the work.

We thank the staff supporting the SET-NET work, including Breanne Anderson with the Arizona Department of Health Services, Tonia Ruddock, J. Michael Bryan, Jerusha Barton, Cyndi Carpentieri, Michael Andrews, Nehali Shah, Amaris Beatty, Andre’a Wilson, Jamil Williams, Carol Zafiratos, Liz Burkhardt, Latasha Terry, and Mildred Banks with the Georgia Department of Public Health, Genna Owens with the Michigan Department of Health and Human Services, Nadia Thomas, Kaylee Mahoney, and Ciarra McFarland with the New York State Department of Health, Rachel Amiya, Zacharie Bakwa, and Caro Johnson with the Washington State Department of Health, and New Jersey Department of Health, for data collection, reporting, and partnership in this important work. This project was also supported in part by an appointment to the Research Participation Program at the Centers for Disease Control and Prevention administered by the Oak Ridge Institute for Science and Education through an interagency agreement between the U.S. Department of Energy and the Centers for Disease Control and Prevention.

